# Hidden overweight and adverse birth outcomes among Asian mothers in the United States, 2016-2023: a population-based repeated cross-sectional study

**DOI:** 10.64898/2026.11.04.26367328

**Authors:** Jabin Tasnim Jahan, Sadia Afrose

**Affiliations:** Department of Epidemiology and Biostatistics, University at Albany, State University of New York, Albany, New York, USA; Chandpur Medical College, Chittagong Medical University, Chittagong, Bangladesh

**Keywords:** Body Mass Index, Asian, Overweight, Hypertension, Pregnancy-Induced, Health Status Disparities

## Abstract

**Background:** Standard body mass index (BMI) categories may understate risk in Asian individuals. Lower Asian action points exist, but their relevance to birth outcomes is unclear.

**Objectives:** To estimate the proportion of normal-weight Asian mothers with BMI 23.0–24.9 kg/m^2^ and its association with birth outcomes, overall and by Asian subgroup, and to compare Asian and White mothers.

**Methods:** Repeated cross-sectional study of US singleton live births, 2016–2023 (National Vital Statistics System). Outcomes were hypertensive disorders, caesarean delivery, preterm birth, and low birthweight. Modified Poisson regression, adjusted for confounders and Asian subgroup, estimated risk ratios (RRs), standardized risks, and risk differences (RDs). We tested for a change in slope at 23.0 kg/m^2^, subgroup heterogeneity, and race × BMI interaction.

**Results:** Among 1,758,116 non-Hispanic Asian mothers, 314,510 (17.9%) had BMI 23.0–24.9 kg/m^2^, 30.4% of normal-weight mothers. Compared with BMI 18.5–22.9, standardized risk of hypertensive disorders was 4.5% versus 3.2% (RD 1.4 percentage points, 95% CI 1.3, 1.5; RR 1.44, 95% CI 1.40, 1.47), and of caesarean delivery 32.4% versus 28.6% (RR 1.14, 95% CI 1.13, 1.14). Preterm birth risk was 7.2% versus 6.6% (RR 1.08, 95% CI 1.06, 1.10); low birthweight risk was not higher (7.1% versus 7.5%; RR 0.95, 95% CI 0.93, 0.97). The hypertensive-disorder slope showed no evidence of change at 23.0 (ratio of per-unit RRs 0.99, 95% CI 0.98, 1.01); the hypertensive-disorder RR ranged from 1.24 (95% CI 1.18, 1.29) among Asian Indian to 1.70 (95% CI 1.60, 1.81) among Chinese mothers. BMI–outcome associations differed between Asian and White mothers.

**Conclusions:** Nearly one in three normal-weight Asian mothers had BMI 23.0–24.9 kg/m^2^, associated with higher risks of hypertensive disorders and caesarean delivery but not low birthweight. The hypertensive-disorder gradient showed no evidence of change at 23.0 kg/m^2^, and associations varied across Asian subgroups.

**Synopsis:** *Study question:* Is prepregnancy BMI 23.0–24.9 kg/m^2^, classified as normal weight under standard criteria, associated with adverse birth outcomes among Asian mothers, and does this vary across Asian subgroups?

*What’s already known:* Standard BMI categories were derived mainly from European populations and may understate risk in Asian individuals. Prior studies of Asian-specific thresholds examined single outcomes or restricted samples.

*What this study adds:* Among 1.76 million US births to Asian mothers, nearly one in three normal-weight mothers had BMI 23.0–24.9 kg/m^2^. This range was associated with higher risks of hypertensive disorders and caesarean delivery, but not low birthweight. Hypertensive-disorder risk rose steadily, with no evidence of a slope change at 23.0, and associations varied across Asian subgroups.

## BACKGROUND

Prepregnancy body mass index (BMI) is a well-established determinant of adverse birth outcomes. Higher maternal BMI is associated with increased risk of hypertensive disorders of pregnancy, with a monotonic dose-response relationship across mild and severe subtypes.^1^ Overweight and obesity are associated with elevated risks of caesarean delivery^2^ and of induced and early preterm birth,^3^ while maternal underweight independently predicts preterm birth and low birthweight.^4^

Standard WHO BMI classifications derive predominantly from European populations^5^ and may inadequately characterise adiposity-related risk in Asian individuals.^6^ At any given BMI, Asian populations carry a higher percentage of body fat than White populations of the same age and sex,^7^ with greater abdominal adiposity despite lower body mass.^6^ A WHO expert consultation concluded that 25 kg/m^2^ does not adequately identify cardiometabolic risk in Asian populations and proposed 23.0 and 27.5 kg/m^2^ as public health action points.^6^ US birth-certificate surveillance typically classifies BMI using standard WHO categories,^8^ and Asian American health data are often omitted, aggregated, or inappropriately extrapolated.^9^

Asian Americans are among the fastest-growing racial and ethnic populations in the United States.^10^ They experience documented perinatal inequities, including the highest gestational diabetes rate among the largest racial and ethnic groups^11^ and elevated risk of adverse maternal outcomes compared with White mothers.^12,13^ Several NVSS studies have examined prepregnancy BMI in these disparities, but each has covered only one piece: analyses have grouped Asian mothers with other races under standard thresholds,^14^ applied Asian-specific thresholds to a single outcome such as preterm birth^15^ or gestational diabetes,^16^ or examined hypertensive disorders or obesity prevalence without a multi-outcome analysis.^17,18^ Studies in single states and health systems show that Asian-specific thresholds classify many more Asian women as overweight^19^ and substantially increase the population-attributable fraction of overweight and obesity for gestational diabetes.^20^ Evidence on whether lower thresholds improve risk identification is inconsistent: in a UK cohort, the South Asian cut-off increased obesity prevalence but did not improve prediction of adverse perinatal outcomes.^21^

Using NVSS natality data from 2016 to 2023, this study aimed to: (a) quantify the proportion of standard normal-weight non-Hispanic Asian mothers with BMI 23.0–24.9 kg/m^2^, overall, by maternal age, and among Asian Indian mothers; (b) estimate the association between Asian-specific BMI categories and hypertensive disorders, caesarean delivery, preterm birth, and low birthweight among non-Hispanic Asian mothers, and assess whether these associations vary across detailed Asian subgroups; and (c) as a secondary aim, compare Asian and White mothers and assess whether the BMI–outcome association differs between them.

## METHODS

### Study design and data source

We conducted a population-based repeated cross-sectional analysis of singleton live births in the United States from 2016 (the first year of full national implementation of the 2003 revised birth certificate) through 2023, using the National Vital Statistics System (NVSS) Natality Public-Use Files.^22,23^ This study followed the STROBE guideline.^24^ The data are publicly available and de-identified, so institutional review board approval was not required.

### Study population

From 29,950,846 births occurring in the United States, including births to non-US residents, we excluded multiple gestations, records with missing or implausible prepregnancy BMI (outside 13.0–69.9 kg/m^2^), and records with missing maternal age, race/ethnicity, or nativity (Figure 1). Excluded records had higher proportions of low birthweight and caesarean delivery, mainly because they included multiple gestations (eTable 2). The primary analysis examined BMI and outcomes among single-race non-Hispanic Asian mothers (n = 1,758,116). Single-race non-Hispanic White mothers (n = 14,486,078) were the comparison group for secondary analyses, because standard BMI thresholds were derived largely from populations of European ancestry. Asian mothers were further classified by the NVSS detailed race recode as Asian Indian, Chinese, Filipino, Japanese, Korean, Vietnamese, or other Asian. Nativity was classified as US-born or foreign-born. Race and ethnicity were treated as social constructs, used to describe perinatal inequities.

**Figure 1.**
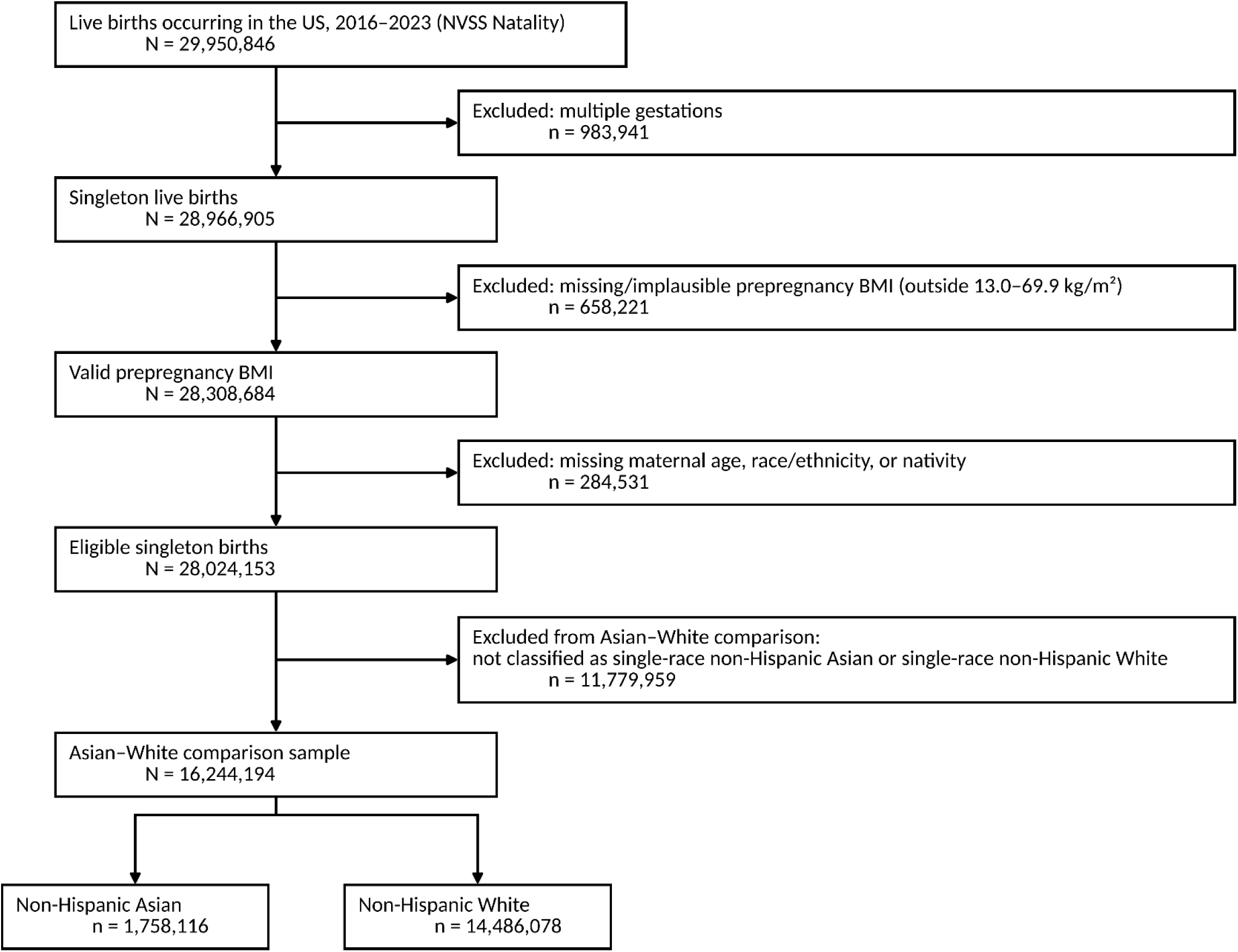
Flow diagram of study population selection, National Vital Statistics System Natality, United States, 2016–2023. The starting count includes all live births occurring in the United States, including births to non-US residents.

### Exposure: BMI classification

Prepregnancy BMI was calculated from self-reported height and weight and classified using standard WHO categories^25^ and study-defined categories based on the WHO public health action points for Asian populations (23.0 and 27.5 kg/m^2^).^6^ For analyses among Asian mothers, we used five categories: underweight (<18.5), normal weight (18.5–22.9, reference), hidden overweight (23.0–24.9; normal under standard criteria but above the first Asian action point), visible overweight (25.0–27.4), and obese (≥27.5 kg/m^2^). Underweight was kept as a separate category because it is associated with preterm birth and low birthweight.^4^

### Outcomes

Outcomes were hypertensive disorders of pregnancy (gestational hypertension, including pre-eclampsia, or eclampsia; chronic hypertension alone was not counted as an outcome, and mothers with chronic hypertension were retained), caesarean delivery, preterm birth (<37 weeks, obstetric estimate), and low birthweight (<2500 g).

### Covariates

Models adjusted for maternal age category (reference 25–29 years), education, marital status, live birth order, prenatal care timing, payment source, smoking, nativity, and birth year (reference 2016; to account for secular trends in hypertensive disorders^26^), selected using a directed acyclic graph (eFigure 1). Infant sex was included as a precision covariate. Gestational diabetes and gestational weight gain were considered mediators and were not adjusted for. Models restricted to Asian mothers also adjusted for detailed Asian subgroup. Unknown categories of education, prenatal care, payment source, and birth order were kept as separate levels.

## Statistical analysis

We estimated risk ratios (RRs) using modified Poisson regression with a log link,^27^ fitted in SAS 9.4 PROC GLIMMIX with empirical (sandwich) variance.^28^ Figures were produced in R 4.6.0 with ggplot2.^29,30^ Estimates are covariate-adjusted associations, not causal effects.

We described the proportion of standard-normal-weight Asian mothers with BMI 23.0–24.9 kg/m^2^, overall, among Asian Indian mothers, and by maternal age.

Among Asian mothers, we estimated RRs for each BMI category relative to 18.5–22.9 kg/m^2^. To show absolute risks, we calculated standardized risks for each BMI category by marginal standardization: we set every mother’s BMI category to each level in turn and averaged the model-predicted risks.^31^ Risk differences and RRs were derived from these standardized risks, with 95% CIs from the delta method.

To test whether risk changed at 23.0 kg/m^2^, we restricted the sample to Asian mothers with BMI 18.5–27.4 kg/m^2^ and fitted models with continuous BMI and a hinge term, max(BMI − 23.0, 0). Slopes were estimated on the log-risk scale. The exponentiated hinge coefficient is the ratio of the RR per 1 kg/m^2^ above versus below 23.0, and a value of 1 indicates no change in slope (eTable 4).

To assess variation across Asian subgroups, we fitted the five-category BMI model separately within each subgroup. Heterogeneity of the BMI 23.0–24.9 RR across subgroups was tested with a 6-degree-of-freedom Wald test of BMI category by subgroup interaction terms, in mothers with BMI 18.5–24.9 kg/m^2^ (Table 4).

In secondary analyses, we estimated adjusted Asian versus White RRs for each outcome, adjusting for standard BMI category. To test whether the BMI–outcome association differed between Asian and White mothers, we modelled BMI with a restricted cubic spline^32^ with 5 knots at the 5th, 27.5th, 50th, 72.5th, and 95th percentiles,^33^ with race × spline interaction terms, tested with a 4-degree-of-freedom Wald test. From these models, we estimated Asian versus White RRs at selected BMI values and plotted RRs across BMI 17–40 kg/m^2^ relative to White mothers at 25 kg/m^2^ (eTable 5; eFigure 2). We repeated the interaction tests within US-born and foreign-born mothers and after excluding 2020–2021, and we tested a race × BMI × nativity interaction (4 degrees of freedom).

### Missing data

Marital status was missing for 23.6% of Asian and 5.3% of White mothers, mainly because California has not reported marital status since 2017.^22^ California accounts for about 26% of US Asian births.^34^ All other covariates were missing for <0.2% of records. Primary analyses were complete-case. We repeated the main Asian BMI models with missing marital status as a separate category and with marital status omitted, which retains California births. For hypertensive disorders, we also used multiple imputation by fully conditional specification^35^ (50 datasets), with the outcome, BMI category, all covariates, and Asian subgroup in the imputation model, and pooled the results using Rubin’s rules^36^ (eTable 6).

### Sensitivity analyses

For hypertensive disorders, we repeated the main Asian BMI model after excluding eclampsia from the outcome, and restricted to births in 2022–2023. We calculated E-values for the hypertensive-disorder RRs at BMI 23.0–24.9 and ≥27.5 kg/m^2^ (eTable 6).^37^

## RESULTS

### Analytic sample

After excluding multiple gestations (n = 983,941), records with missing or implausible prepregnancy BMI (n = 658,221), and records with missing maternal age, race/ethnicity, or nativity (n = 284,531), the analytic sample included 28,024,153 singleton live births (Figure 1). Of these, 1,758,116 (6.3%) were to non-Hispanic Asian mothers and 14,486,078 (51.7%) to non-Hispanic White mothers. Complete-case regression samples included 1,339,153 to 1,340,254 Asian mothers and 14,990,774 to 15,004,335 Asian and White mothers, depending on the outcome. Most exclusions from these samples were due to missing marital status, which was not recorded for 23.6% of Asian and 5.3% of White mothers. Sample sizes and event counts for each model are shown in eTable 3.

### Sample characteristics

Compared with White mothers, Asian mothers were older (mean 32.1 versus 29.6 years), had lower prepregnancy BMI (mean 24.1 versus 26.9 kg/m^2^), were more often foreign-born (79.2% versus 6.6%) and college-educated (65.2% versus 43.5%), and smoked less (0.4% versus 8.0%) (Table 1). Crude prevalences among Asian and White mothers were 5.0% and 8.5% for hypertensive disorders, 31.9% and 29.4% for caesarean delivery, 7.3% and 7.3% for preterm birth, and 7.3% and 5.3% for low birthweight (Table 2).

**Table 1.** Maternal characteristics of non-Hispanic Asian and non-Hispanic White mothers, United States singleton live births, 2016–2023.

| Characteristic | Asian n (%) [N = 1,758,116] | White n (%) [N = 14,486,078] | \|d\| |
|---|---|---|---|
| Maternal age (years) |  |  |  |
| Mean (SD) | 32.1 (4.8) | 29.6 (5.5) | 0.48 |
| <20 | 10,006 (0.6) | 460,169 (3.2) | 0.19 |
| 20–24 | 99,467 (5.7) | 2,370,803 (16.4) | 0.35 |
| 25–29 | 396,986 (22.6) | 4,210,937 (29.1) | 0.15 |
| 30–34 | 712,512 (40.5) | 4,685,024 (32.3) | 0.17 |
| 35–39 | 437,155 (24.9) | 2,297,877 (15.9) | 0.22 |
| ≥40 | 101,990 (5.8) | 461,268 (3.2) | 0.13 |
| Prepregnancy BMI (kg/m<sup>2</sup>) |  |  |  |
| Mean (SD) | 24.1 (4.6) | 26.9 (6.7) | 0.49 |
| Foreign-born |  |  |  |
| Yes | 1,392,932 (79.2) | 957,989 (6.6) | 2.16 |
| Maternal education |  |  |  |
| Less than high school | 102,388 (5.8) | 972,206 (6.7) | 0.04 |
| High school diploma/GED | 205,155 (11.7) | 3,075,092 (21.2) | 0.26 |
| Some college | 277,492 (15.8) | 4,059,058 (28.0) | 0.30 |
| Bachelor's degree | 607,155 (34.5) | 3,931,896 (27.1) | 0.16 |
| Graduate degree | 539,105 (30.7) | 2,375,038 (16.4) | 0.34 |
| Unknown | 26,821 (1.5) | 72,788 (0.5) | 0.10 |
| Marital status<sup>a</sup> |  |  |  |
| Married | 1,184,455 (67.4) | 9,823,576 (67.8) | 0.01 |
| Unmarried | 158,837 (9.0) | 3,890,048 (26.9) | 0.48 |
| Missing | 414,824 (23.6) | 772,454 (5.3) | 0.54 |
| Live birth order |  |  |  |
| 1st birth | 808,327 (46.0) | 5,861,577 (40.5) | 0.11 |
| 2nd birth | 648,708 (36.9) | 4,772,442 (32.9) | 0.08 |
| 3rd birth | 202,808 (11.5) | 2,304,840 (15.9) | 0.13 |
| ≥4th birth | 95,225 (5.4) | 1,514,876 (10.5) | 0.19 |
| Unknown | 3,048 (0.2) | 32,343 (0.2) | 0.01 |
| Prenatal care timing |  |  |  |
| 1st trimester | 1,416,977 (80.6) | 11,808,109 (81.5) | 0.02 |
| 2nd trimester | 230,045 (13.1) | 1,827,483 (12.6) | 0.01 |
| 3rd trimester | 65,506 (3.7) | 461,287 (3.2) | 0.03 |
| No prenatal care | 15,337 (0.9) | 161,614 (1.1) | 0.03 |
| Unknown | 30,251 (1.7) | 227,585 (1.6) | 0.01 |
| Payment source |  |  |  |
| Private insurance | 1,210,238 (68.8) | 9,208,692 (63.6) | 0.11 |
| Medicaid | 411,323 (23.4) | 4,204,824 (29.0) | 0.13 |
| Self-pay | 77,207 (4.4) | 479,399 (3.3) | 0.06 |
| Other | 51,406 (2.9) | 494,209 (3.4) | 0.03 |
| Unknown | 7,942 (0.5) | 98,954 (0.7) | 0.03 |
| Smoking during pregnancy |  |  |  |
| Yes | 6,988 (0.4) | 1,159,401 (8.0) | 0.39 |

<sup>a</sup> Missing marital status is shown as a separate category for descriptive purposes because missingness was substantial and differed by race/ethnicity.
Percentages use race/ethnicity-specific denominators and may not sum to 100 because of rounding. Unknown categories for education, prenatal care, payment source, and birth order were retained as separate levels in regression models. \|d\|, absolute standardized difference; BMI, body mass index; SD, standard deviation.

**Table 2.** Crude and adjusted risk ratios for adverse birth outcomes among non-Hispanic Asian versus non-Hispanic White mothers, United States singleton births, 2016–2023.

| **Outcome** | **Asian, n/N (%)** | **White, n/N (%)** | **Crude RR (95% CI)** | **Adjusted RR (95% CI)<sup>a</sup>** |
|---|---|---|---|---|
| Hypertensive disorders | 88,355/1,757,124 (5.0) | 1,234,609/14,472,881 (8.5) | 0.59 (0.59, 0.59) | 0.91 (0.90, 0.92) |
| Caesarean delivery | 560,533/1,756,636 (31.9) | 4,254,637/14,467,419 (29.4) | 1.09 (1.08, 1.09) | 1.08 (1.07, 1.08) |
| Preterm birth | 129,049/1,757,780 (7.3) | 1,058,385/14,480,377 (7.3) | 1.00 (1.00, 1.01) | 1.26 (1.25, 1.27) |
| Low birthweight | 128,734/1,757,413 (7.3) | 762,177/14,478,840 (5.3) | 1.39 (1.38, 1.40) | 1.65 (1.63, 1.66) |

<sup>a</sup> Referent: non-Hispanic White mothers. Modified Poisson regression with robust variance, adjusted for standard WHO BMI category, maternal age category, infant sex, nativity, education, marital status, live birth order, prenatal care timing, payment source, smoking, and birth year. Complete-case analysis; missing marital status accounted for most exclusions (414,824 Asian [23.6%] and 772,454 White [5.3%] mothers). With missing marital status as a separate category, adjusted RRs were 0.93 (95% CI 0.93, 0.94) for hypertensive disorders and 1.05 (95% CI 1.05, 1.06) for caesarean delivery; preterm birth and low birthweight estimates were unchanged. These RRs estimate the overall Asian–White association conditional on standard BMI category and covariates. Because the association varied with BMI (restricted cubic spline race × BMI interaction, all P < 0.0001), they should be read with the BMI-specific estimates in the text, eTable 5, and eFigure 2.
CI, confidence interval; RR, risk ratio.

### Classification under Asian-specific thresholds

Among Asian mothers, 314,510 (17.9%) had BMI 23.0–24.9 kg/m^2^, representing 30.4% of those classified as normal weight under standard criteria. This proportion increased with maternal age, from 29.1% at ages 20–24 to 31.8% at 40 or older, and was higher among Asian Indian mothers (38.5%; 32.9% to 41.1% across age groups) (eFigure 3; eTable 1).

### BMI and adverse outcomes among Asian mothers

Among Asian mothers, risk of hypertensive disorders and caesarean delivery increased with BMI (Table 3; Figure 2). Compared with BMI 18.5–22.9, the standardized risk of hypertensive disorders at BMI 23.0–24.9 was 4.5% versus 3.2% (risk difference [RD] 1.4 percentage points, 95% CI 1.3, 1.5; risk ratio [RR] 1.44, 95% CI 1.40, 1.47), rising to 8.8% at BMI ≥27.5 (RR 2.79, 95% CI 2.73, 2.85). For caesarean delivery, the standardized risk at BMI 23.0–24.9 was 32.4% versus 28.6% (RD 3.9, 95% CI 3.7, 4.1; RR 1.14, 95% CI 1.13, 1.14), rising to 40.4% at BMI ≥27.5 (RR 1.41, 95% CI 1.41, 1.42).

**Figure 2.**
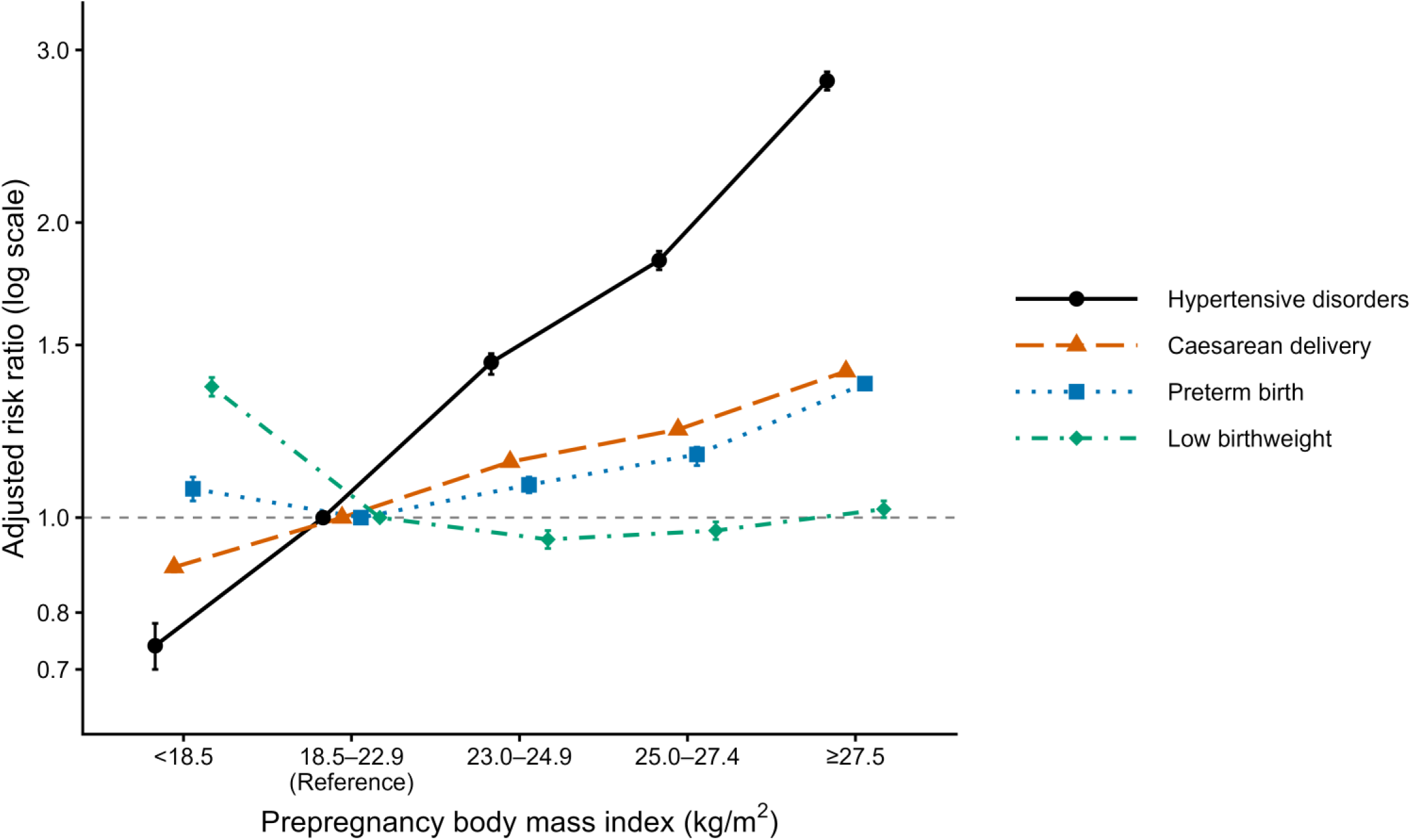
Adjusted risk ratios for adverse birth outcomes across Asian-specific prepregnancy body mass index (BMI) categories among non-Hispanic Asian mothers, National Vital Statistics System, 2016–2023. Risk ratios were estimated using modified Poisson regression with robust variance, adjusted for maternal age, infant sex, nativity, education, marital status, live birth order, prenatal care timing, payment source, smoking, birth year, and detailed Asian subgroup. BMI 18.5–22.9 kg/m^2^ was the reference category. Vertical bars indicate 95% confidence intervals; some intervals are narrower than the plotting symbols. The y-axis is on a logarithmic scale.

**Table 3.** Adverse birth outcomes across Asian-specific prepregnancy BMI categories among non-Hispanic Asian mothers, United States singleton births, 2016–2023 (referent: 18.5–22.9 kg/m^2^).

| BMI category (kg/m<sup>2</sup>) | n/N (%)<sup>a</sup> | Standardized risk, % (95% CI)<sup>b</sup> | Risk difference, percentage points (95% CI)<sup>b</sup> | Adjusted RR (95% CI)<sup>b</sup> |
|---|---|---|---|---|
| Hypertensive disorders |  |  |  |  |
| Underweight <18.5 | 1,783/83,964 (2.1) | 2.3 (2.2, 2.4) | −0.8 (−0.9, −0.7) | 0.74 (0.70, 0.78) |
| Normal 18.5–22.9 | 16,590/538,655 (3.1) | 3.2 (3.1, 3.2) | 0.00 (Reference) | 1.00 (Reference) |
| Hidden 23.0–24.9 | 10,996/241,107 (4.6) | 4.5 (4.5, 4.6) | 1.4 (1.3, 1.5) | 1.44 (1.40, 1.47) |
| Visible overweight 25.0–27.4 | 12,124/206,617 (5.9) | 5.8 (5.7, 5.9) | 2.6 (2.5, 2.7) | 1.83 (1.79, 1.87) |
| Obese ≥27.5 | 25,161/269,286 (9.3) | 8.8 (8.7, 8.9) | 5.7 (5.5, 5.8) | 2.79 (2.73, 2.85) |
| Caesarean delivery |  |  |  |  |
| Underweight <18.5 | 19,750/83,937 (23.5) | 25.3 (25.0, 25.6) | −3.2 (−3.6, −2.9) | 0.89 (0.88, 0.90) |
| Normal 18.5–22.9 | 151,331/538,459 (28.1) | 28.6 (28.4, 28.7) | 0.00 (Reference) | 1.00 (Reference) |
| Hidden 23.0–24.9 | 79,827/241,028 (33.1) | 32.4 (32.3, 32.6) | 3.9 (3.7, 4.1) | 1.14 (1.13, 1.14) |
| Visible overweight 25.0–27.4 | 74,307/206,540 (36.0) | 35.0 (34.8, 35.2) | 6.5 (6.2, 6.7) | 1.23 (1.22, 1.23) |
| Obese ≥27.5 | 110,066/269,189 (40.9) | 40.4 (40.2, 40.6) | 11.8 (11.6, 12.1) | 1.41 (1.41, 1.42) |
| Preterm birth |  |  |  |  |
| Underweight <18.5 | 5,535/83,997 (6.6) | 7.1 (6.9, 7.3) | 0.5 (0.3, 0.7) | 1.07 (1.04, 1.10) |
| Normal 18.5–22.9 | 34,629/538,917 (6.4) | 6.6 (6.6, 6.7) | 0.00 (Reference) | 1.00 (Reference) |
| Hidden 23.0–24.9 | 17,389/241,249 (7.2) | 7.2 (7.1, 7.3) | 0.5 (0.4, 0.6) | 1.08 (1.06, 1.10) |
| Visible overweight 25.0–27.4 | 16,291/206,726 (7.9) | 7.7 (7.6, 7.8) | 1.0 (0.9, 1.2) | 1.16 (1.13, 1.18) |
| Obese ≥27.5 | 26,000/269,365 (9.7) | 9.1 (9.0, 9.2) | 2.5 (2.3, 2.6) | 1.37 (1.35, 1.39) |
| Low birthweight |  |  |  |  |
| Underweight <18.5 | 7,702/83,980 (9.2) | 10.2 (9.9, 10.4) | 2.7 (2.5, 2.9) | 1.36 (1.33, 1.39) |
| Normal 18.5–22.9 | 38,098/538,822 (7.1) | 7.5 (7.4, 7.6) | 0.00 (Reference) | 1.00 (Reference) |
| Hidden 23.0–24.9 | 17,423/241,184 (7.2) | 7.1 (7.0, 7.2) | −0.4 (−0.5, −0.3) | 0.95 (0.93, 0.97) |
| Visible overweight 25.0–27.4 | 15,695/206,661 (7.6) | 7.2 (7.1, 7.4) | −0.2 (−0.4, −0.1) | 0.97 (0.95, 0.99) |
| Obese ≥27.5 | 22,315/269,264 (8.3) | 7.6 (7.5, 7.7) | 0.2 (0.0, 0.3) | 1.02 (1.00, 1.04) |

<sup>a</sup> Crude counts in the complete-case regression sample (1,339,153 to 1,340,254 mothers across outcomes).
<sup>b</sup> Modified Poisson regression with robust variance, adjusted for detailed Asian subgroup, maternal age category, infant sex, nativity, education, marital status, live birth order, prenatal care timing, payment source, smoking, and birth year. Standardized risks are model-predicted risks averaged over the complete-case Asian sample with BMI set to each category (marginal standardization). CIs for standardized risks, risk differences, and RRs were derived by the delta method. Adjusted RRs are standardized risk ratios; both RDs and RRs were calculated from unrounded standardized risks.
CI, confidence interval; RD, risk difference; RR, risk ratio.

**Table 4.** Risk ratios for BMI 23.0–24.9 versus 18.5–22.9 kg/m^2^ by detailed Asian subgroup, United States singleton births, 2016–2023.

| Subgroup | Model N, hypertensive disorders<sup>a</sup> | BMI 23.0–24.9 among standard normal weight, %<sup>b</sup> | Hypertensive disorders RR (95% CI)<sup>c</sup> | Caesarean delivery RR (95% CI)<sup>c</sup> | Preterm birth RR (95% CI)<sup>c</sup> | Low birthweight RR (95% CI)<sup>c</sup> |
|---|---|---|---|---|---|---|
| Asian Indian | 454,578 | 38.5 | 1.24 (1.18, 1.29) | 1.10 (1.09, 1.11) | 1.03 (1.00, 1.06) | 0.92 (0.90, 0.95) |
| Chinese | 244,793 | 21.7 | 1.70 (1.60, 1.81) | 1.16 (1.14, 1.19) | 1.13 (1.08, 1.18) | 0.98 (0.93, 1.03) |
| Filipino | 144,273 | 35.8 | 1.41 (1.34, 1.49) | 1.11 (1.09, 1.13) | 1.12 (1.07, 1.17) | 1.00 (0.95, 1.05) |
| Japanese | 31,233 | 21.4 | 1.65 (1.41, 1.93) | 1.16 (1.10, 1.23) | 1.11 (0.97, 1.26) | 0.81 (0.70, 0.93) |
| Korean | 72,715 | 24.5 | 1.55 (1.41, 1.71) | 1.20 (1.16, 1.24) | 1.13 (1.04, 1.23) | 0.92 (0.84, 1.01) |
| Vietnamese | 106,828 | 23.0 | 1.68 (1.54, 1.84) | 1.13 (1.10, 1.16) | 1.12 (1.05, 1.19) | 1.01 (0.95, 1.08) |
| Other Asian | 285,209 | 34.9 | 1.33 (1.26, 1.40) | 1.14 (1.12, 1.16) | 1.03 (0.99, 1.07) | 0.93 (0.89, 0.96) |
| All Asian, subgroup-adjusted | 1,339,629 | 30.4 | 1.44 (1.40, 1.47) | 1.14 (1.13, 1.14) | 1.08 (1.06, 1.10) | 0.95 (0.93, 0.97) |
| Heterogeneity χ² (6 df), P<sup>d</sup> |  |  | 121.5, <0.0001 | 70.9, <0.0001 | 25.7, 0.0002 | 24.7, 0.0004 |

<sup>a</sup> Births in the complete-case hypertensive-disorder model. Ns for the other outcomes differed by <0.1%.
<sup>b</sup> Proportion of births to Asian mothers with BMI 18.5–24.9 kg/m<sup>2</sup> (normal weight under standard WHO criteria) whose BMI was 23.0–24.9 kg/m<sup>2</sup>, calculated in the full eligible sample before complete-case exclusions. Model sample sizes reflect complete-case analyses.
<sup>c</sup> Modified Poisson regression with robust variance, fitted separately within each subgroup and adjusted for the covariates listed in Table 3 (except subgroup). Complete-case analysis. Models include all five BMI categories; only the BMI 23.0–24.9 contrast is shown.
<sup>d</sup> Joint Wald test of the BMI category (23.0–24.9 versus 18.5–22.9) by subgroup interaction terms, in a model restricted to mothers with BMI 18.5–24.9 kg/m<sup>2</sup>.
Subgroups were identified from the NVSS detailed maternal race recode. CI, confidence interval; RR, risk ratio.

Associations with preterm birth were weaker (BMI 23.0–24.9: RD 0.5, 95% CI 0.4, 0.6; RR 1.08, 95% CI 1.06, 1.10). Low birthweight risk was slightly lower at BMI 23.0–24.9 than in the reference group (RD −0.4, 95% CI −0.5, −0.3; RR 0.95, 95% CI 0.93, 0.97) and highest among underweight mothers (RR 1.36, 95% CI 1.33, 1.39). In the same sample and model without adjustment for Asian subgroup, the low birthweight RR at BMI 23.0–24.9 was 1.03 (95% CI 1.02, 1.05).

On the log-risk scale, there was no evidence that the RR per 1 kg/m^2^ changed at 23.0 kg/m^2^ for hypertensive disorders (ratio of per-unit RRs above versus below 23.0: 0.991, 95% CI 0.976, 1.006; P = 0.24). The ratio was 0.990 (95% CI 0.985, 0.994) for caesarean delivery, 1.025 (95% CI 1.014, 1.037) for preterm birth, and 1.067 (95% CI 1.055, 1.079) for low birthweight (eTable 4).

### Variation across Asian subgroups

Associations at BMI 23.0–24.9 differed across Asian subgroups for all four outcomes (heterogeneity P ≤ 0.0004) (Table 4). RRs for hypertensive disorders ranged from 1.24 (95% CI 1.18, 1.29) among Asian Indian mothers to 1.70 (95% CI 1.60, 1.81) among Chinese mothers, and RRs for caesarean delivery from 1.10 to 1.20. RRs for preterm birth ranged from 1.03 to 1.13. No subgroup showed a higher risk of low birthweight (RRs 0.81 to 1.01). Asian Indian mothers had the highest proportion with BMI 23.0–24.9 but the weakest associations for hypertensive disorders, caesarean delivery, and preterm birth.

### Asian–White differences

After adjustment, Asian mothers had a lower risk of hypertensive disorders than White mothers (RR 0.91, 95% CI 0.90, 0.92) and higher risks of caesarean delivery (RR 1.08, 95% CI 1.07, 1.08), preterm birth (RR 1.26, 95% CI 1.25, 1.27), and low birthweight (RR 1.65, 95% CI 1.63, 1.66) (Table 2).

These differences varied by BMI for all four outcomes (race × BMI interaction, all P < 0.0001) (eTable 5; eFigure 2). For hypertensive disorders, Asian mothers had lower risk than White mothers at BMI 20 (RR 0.79, 95% CI 0.78, 0.81), with similar risk at BMI 35 (RR 1.02, 95% CI 1.00, 1.03). For caesarean delivery, the Asian excess was largest near BMI 23 (RR 1.13, 95% CI 1.12, 1.13) and reversed at BMI 35 (RR 0.98, 95% CI 0.98, 0.99). For preterm birth, Asian mothers had higher risk from about BMI 20 upward, and the difference grew with BMI (RR 1.07 at BMI 20 and 1.43 at BMI 35). For low birthweight, Asian mothers had higher risk at every BMI (RR 1.38 at BMI 20 and 1.86 at BMI 35, peaking at 1.94 near BMI 27.5). The interaction remained among US-born and foreign-born mothers and after excluding 2020–2021, and it differed by nativity (all P < 0.0001) (eTable 5).

### Sensitivity analyses

The RR for hypertensive disorders at BMI 23.0–24.9 was 1.43 to 1.44 across approaches to missing marital status and after excluding eclampsia, and 1.35 (95% CI 1.29, 1.41) in 2022–2023 alone (eTable 6). The E-value for this estimate was 2.24 (lower confidence limit 2.15). The Asian–White RR for caesarean delivery ranged from 1.04 to 1.08 across approaches to missing marital status; other Asian–White estimates changed little, and none changed direction.

## COMMENT

### Principal findings

About 314,500 non-Hispanic Asian mothers, nearly one in three of those classified as normal weight under standard criteria, had a prepregnancy BMI of 23.0–24.9 kg/m^2^. Compared with BMI 18.5–22.9, this range was associated with higher risks of hypertensive disorders (RR 1.44; 1.4 additional cases per 100 births) and caesarean delivery (RR 1.14; 3.9 per 100), a small increase in preterm birth (RR 1.08), and no increase in low birthweight. For hypertensive disorders, risk rose steadily with BMI, with no evidence of a change in slope at 23.0 kg/m^2^. Associations varied across Asian subgroups, and the BMI–outcome association differed between Asian and White mothers for all four outcomes.

Prior NVSS-based analyses applied Asian-specific thresholds to single outcomes or restricted samples.^14–18^ A Korean cohort showed obstetric risk rising at BMI ≥23.0 kg/m^2^,^38^ but did not quantify reclassification or compare Asian and White mothers. The only multi-outcome study of South Asian-specific thresholds found largely linear associations without improved risk prediction, but was limited to one UK population.^21^ The present study examines four outcomes across seven Asian subgroups in national US data.

## Strengths of the study

The study included more than 1.75 million Asian births, allowing subgroup-specific estimates. We reported absolute risks and risk differences alongside RRs, adjusted for detailed Asian subgroup, and kept underweight mothers separate from the reference group.^4^ The hypertensive-disorder estimate was stable across approaches to missing marital status (RR 1.43–1.44), including a model that retained California births, and an unmeasured confounder would need an RR of at least 2.24 with both BMI and the outcome to explain it away. The race × BMI interaction persisted within US-born and foreign-born mothers and after excluding 2020–2021.

## Limitations of the data

Prepregnancy BMI was self-reported, and reporting error varies by race/ethnicity.^39^ If Asian mothers underreport weight, the prevalence and risks for BMI 23.0–24.9 may be underestimated. Because this category spans only 2 kg/m^2^, small reporting errors can move mothers in or out of it. Excluded records had higher adverse-outcome rates, mainly because of multiple gestations, and the same exclusion criteria were applied to all racial groups. Marital status was missing mainly for California births. Multiple imputation assumes data are missing at random, which may not hold for structural missingness, but the model omitting marital status gave the same estimate. The public-use files have no geographic identifier, so the California attribution relies on NCHS documentation.^22^ Birth-certificate reporting of hypertensive disorders has low sensitivity, which may have attenuated associations; birthweight and gestational age are recorded accurately.^40^ Results were similar after excluding eclampsia, which all states reported only from 2022,^22^ and when restricted to 2022–2023. Births to the same mother could not be linked, so births were treated as independent, which may slightly understate standard errors. Prenatal care timing and payment source are recorded during pregnancy; we included them as markers of access to care and socioeconomic position. We could not separate spontaneous from medically indicated preterm birth. The other Asian category is heterogeneous, and multiracial Asian mothers were excluded, which may limit generalisability. NCHS imputes maternal race for about 8% of records,^22^ but misclassification of Asian mothers is likely small. The file includes births to non-US residents (about 0.2% of records). With this sample size, very small differences are statistically significant, so risk differences are a better guide to practical importance. This study describes associations and does not assess predictive performance; future studies should compare discrimination, calibration, and clinical utility of standard and Asian-specific thresholds.

## Interpretation

The higher hypertensive-disorder risk at BMI 23.0–24.9 is biologically plausible. Asian individuals carry more body fat at a given BMI,^6,7^ with more visceral fat,^41^ which promotes insulin resistance, endothelial dysfunction, and inflammation.^42,43^ Insulin resistance develops at lower BMI in Asian populations^44^ and precedes pre-eclampsia.^43^ However, risk rose steadily across BMI, with no evidence of a change at 23.0 kg/m^2^. The 23.0 cut-point appears to mark a point on a continuous gradient rather than a point where risk begins to rise.

Associations differed across subgroups. The hypertensive-disorder RR ranged from 1.24 among Asian Indian mothers to 1.70 among Chinese mothers. Asian Indian mothers had the highest proportion with BMI 23.0–24.9 (38.5%) but the weakest associations, possibly because their metabolic risk is already raised at lower BMI, as shown for type 2 diabetes among South Asian women.^45^ A single Asian-specific cut-point may therefore not fit all subgroups equally. The proportion with BMI 23.0–24.9 also rose with maternal age, when baseline obstetric risk is already higher,^46^ which is relevant because 71.2% of Asian mothers in this study were aged 30 or older.

For caesarean delivery, the excess among Asian mothers narrowed at higher BMI and reversed at BMI 35, consistent with Glazer et al.^47^ Cephalopelvic disproportion may contribute, but data are limited.

Crude preterm birth rates were equal (7.3%), but the adjusted risk was 26% higher among Asian mothers. This is because Asian mothers’ lower-risk profile (less smoking, more education, lower BMI) masks their higher risk in crude comparisons. The small increase in low birthweight at BMI 23.0–24.9 seen in pooled data disappeared after adjustment for subgroup (RR 1.03 to 0.95).

Subgroups with more low-birthweight infants also tended to have higher BMI, which produced the pooled association. Higher low-birthweight rates among Asian mothers overall may partly reflect constitutional smallness. Birthweight differences between White and Asian subgroups range from 115 to 235 g,^48^ which questions whether the 2500-g threshold fits all groups.^49^ Customised growth standards may help distinguish small but healthy infants from growth restriction.^50,51^ Underweight mothers had the highest low-birthweight risk, consistent with prior evidence.^4^ The small increase in preterm birth may reflect adiposity-related inflammation^52^ and medically indicated delivery for hypertensive disorders,^53^ which were 44% more common at BMI 23.0–24.9.

The healthy migrant effect predicts better outcomes among foreign-born mothers,^54^ but the race × BMI interaction persisted in both nativity groups. Foreign-born status has been linked to a 24% higher low-birthweight risk among Asian women, unlike the lower risk among foreign-born Black and Hispanic women.^55^ Nativity modified the race × BMI association most strongly for hypertensive disorders and caesarean delivery.

## Conclusions

About 314,500 Asian mothers entered pregnancy at a BMI classified as normal under standard criteria but associated with higher risks of hypertensive disorders and caesarean delivery. For hypertensive disorders, risk rose steadily with BMI, with no evidence of a change at 23.0 kg/m^2^, and associations varied across Asian subgroups. The WHO and ADA support lower BMI action points for cardiometabolic risk in Asian populations,^6,56^ and these findings suggest the concern extends to some perinatal outcomes. Asian-specific categories could be added to perinatal surveillance with no new data collection, because birth certificates already record prepregnancy BMI. Before clinical use, studies should test whether these categories improve risk prediction and whether subgroup-specific cut-points are needed.

## Supporting information

Supplementary eTables and eFigures

## Data Availability

All data used in this study are publicly available from the National Center for Health Statistics, National Vital Statistics System Natality Public-Use Files, 2016-2023. Analytic code is available on GitHub.

https://www.cdc.gov/nchs/data_access/vitalstatsonline.htm

https://github.com/jtasnimjahan/asian-bmi-birth-outcomes

## Acknowledgements

The authors thank the University at Albany for providing access to SAS software and library resources used in this research.

## Author contributions

JTJ: Conceptualisation, Methodology, Formal analysis, Data curation, Visualisation, Writing – original draft, Writing – review & editing.

SA: Investigation (literature search), Interpretation of study findings, Writing – original draft (supporting), Writing – review & editing.

Both authors approved the final manuscript and agree to be accountable for all aspects of the work.

## Data availability statement

The data underlying this study are publicly available from the National Center for Health Statistics (NCHS) through the National Vital Statistics System (NVSS) Natality Public-Use Files, 2016–2023 (https://www.cdc.gov/nchs/data_access/vitalstatsonline.htm). Analytic code is available at https://github.com/jtasnimjahan/asian-bmi-birth-outcomes

