## Supplementary eTables and eFigures for "Hidden overweight and adverse birth outcomes among Asian mothers in the United States, 2016-2023: a population-based repeated cross-sectional study"

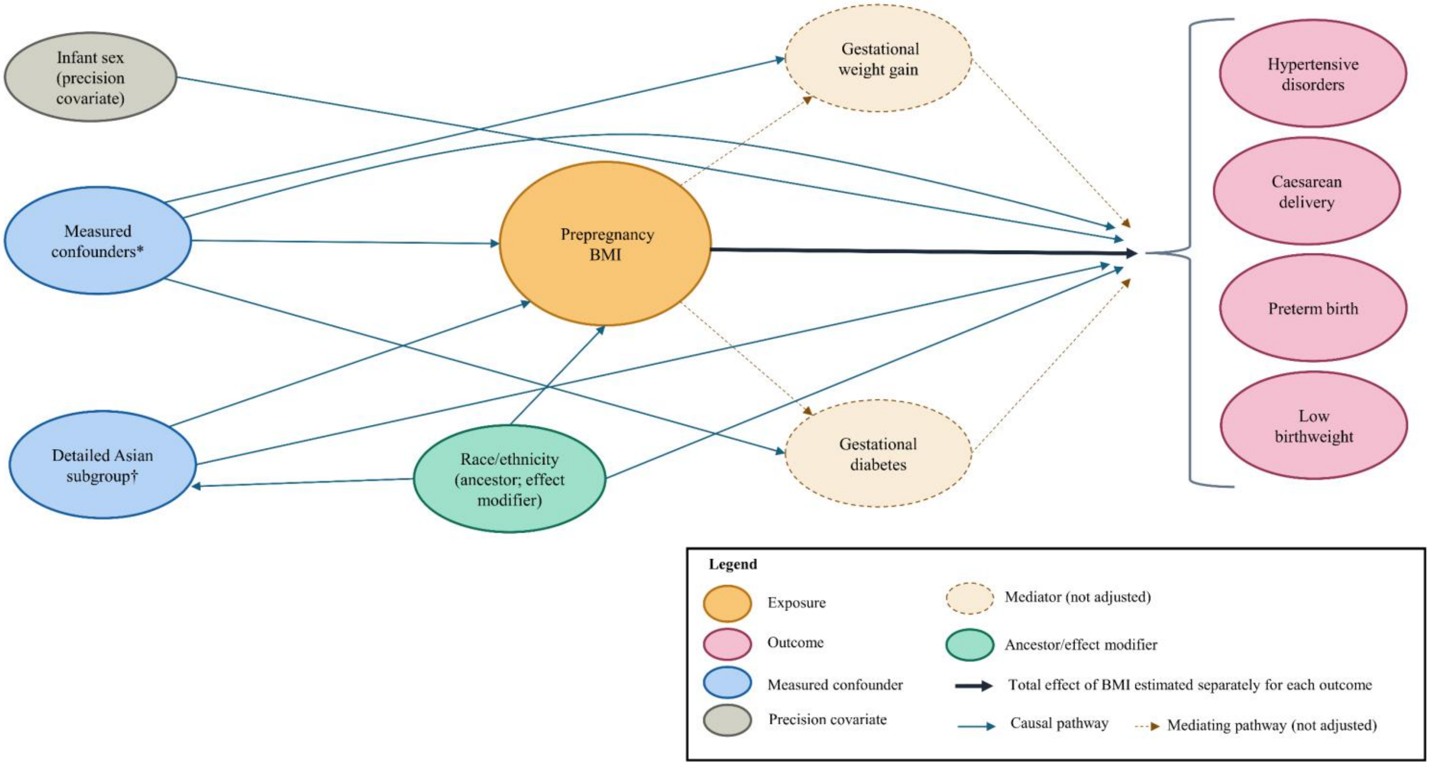


**eFigure 1. Assumed causal structure underlying covariate selection.** Directed acyclic graph for the total effect of prepregnancy body mass index, estimated separately for hypertensive disorders, caesarean delivery, preterm birth, and low birthweight.

*Measured confounders adjusted in all models: maternal age, education, marital status, live birth order, prenatal care timing, payment source, smoking, nativity, and birth year; infant sex was included as a precision covariate. †Detailed Asian subgroup (Asian Indian, Chinese, Filipino, Japanese, Korean, Vietnamese, other Asian) was additionally adjusted in models restricted to Asian mothers. Gestational diabetes mellitus and gestational weight gain were treated as potential mediators and excluded from the primary adjustment set. Arrows pointing to the outcome bracket represent pathways to one or more of the four outcomes shown.

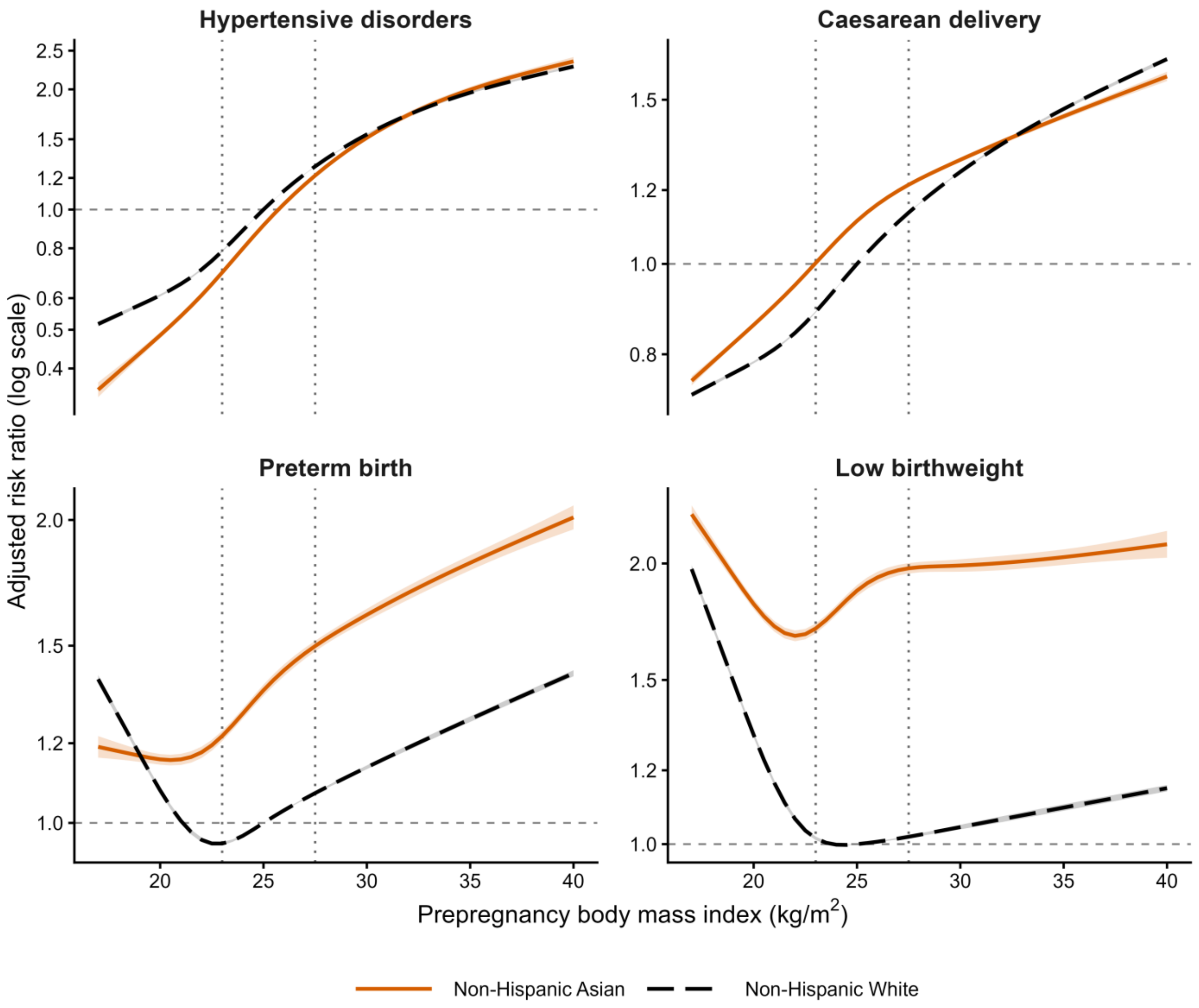


**eFigure 2.** Adjusted risk ratios across prepregnancy body mass index among non-Hispanic Asian and non-Hispanic White mothers, National Vital Statistics System, 2016–2023.

Curves were estimated using modified Poisson regression with robust variance. Body mass index was modelled with a restricted cubic spline (5 knots at the 5th, 27.5th, 50th, 72.5th, and 95th percentiles), with race × spline interaction terms. Risk ratios are relative to non-Hispanic White mothers at a body mass index of 25 kg/m$^2$. Shaded bands indicate 95% confidence intervals; some are narrower than the plotted lines. Dotted vertical lines mark the Asian-specific body mass index action points of 23.0 and 27.5 kg/m$^2$. The y-axis is on a logarithmic scale, and its range differs by panel. Models adjusted for maternal age, education, marital status, live birth order, prenatal care timing, payment source, smoking, nativity, birth year, and infant sex. Interaction tests and risk ratios at selected values of body mass index are in eTable 5.

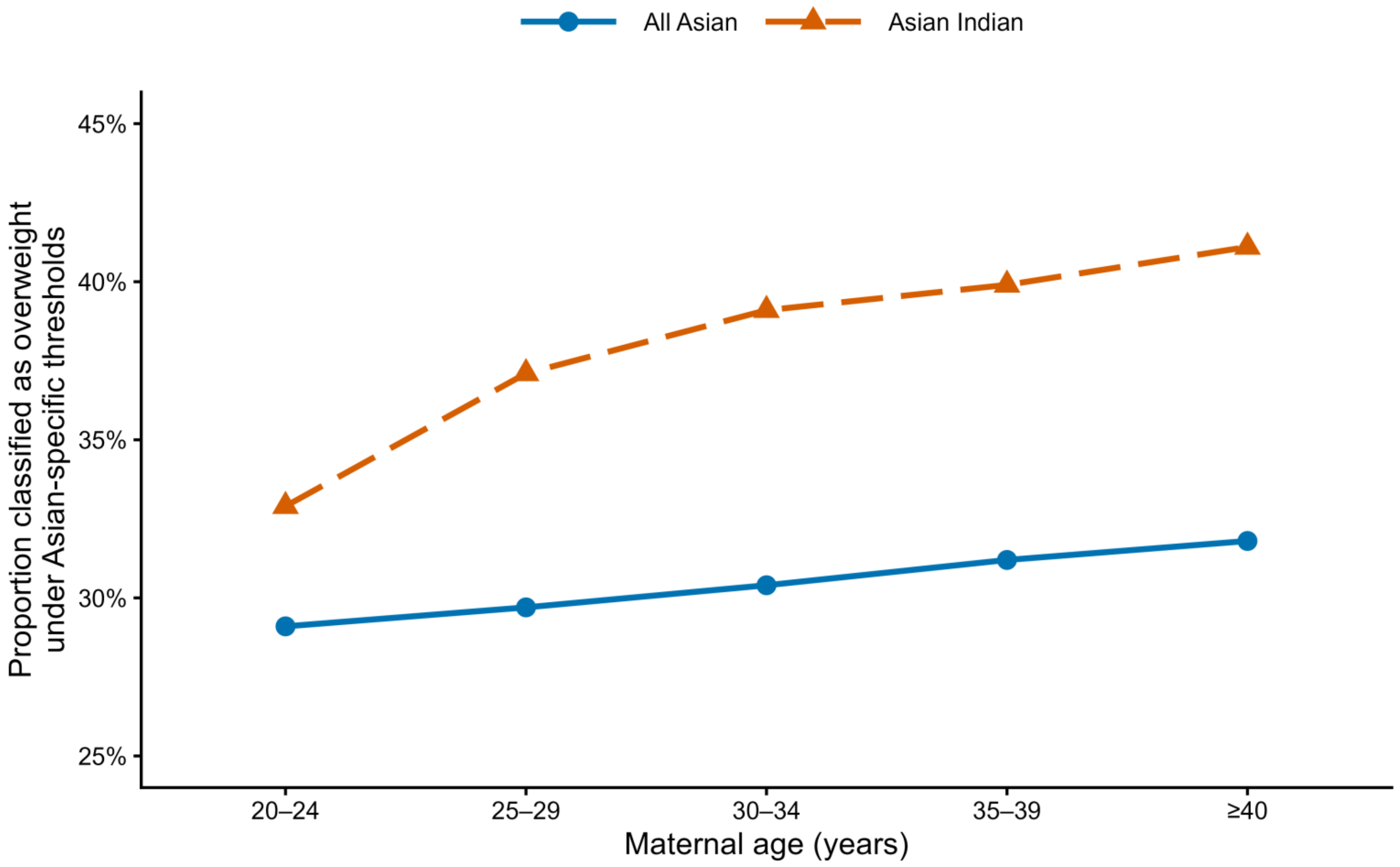


**eFigure 3.** Age-specific proportion of non-Hispanic Asian mothers classified as overweight under Asian-specific body mass index thresholds, United States, 2016–2023.

Proportions were calculated among mothers classified as normal weight under standard criteria (BMI 18.5–24.9 kg/$m^2$). Asian-specific thresholds classify BMI 23.0–24.9 kg/$m^2$ as overweight. Results are shown for all non-Hispanic Asian mothers and Asian Indian mothers. Mothers younger than 20 years (0.6% of Asian births) were omitted from the figure but retained in all analyses.

**eTable 1.** Crude prevalence of adverse birth outcomes by maternal age among Asian Indian mothers in the hidden overweight (BMI 23.0–24.9 kg/m$^2$) and normal-weight groups, United States singleton births, 2016–2023.

| Age band | HD hidden n/N (%) | HD normal n/N (%) | CD hidden n/N (%) | CD normal n/N (%) | PTB hidden n/N (%) | PTB normal n/N (%) | LBW hidden n/N (%) | LBW normal n/N (%) |
|---|---|---|---|---|---|---|---|---|
| 20–24 | 177/4,250 (4.2) | 276/8,672 (3.2) | 1,080/4,248 (25.4) | 1,904/8,670 (22.0) | 261/4,251 (6.1) | 600/8,678 (6.9) | 341/4,249 (8.0) | 930/8,677 (10.7) |
| 25–29 | 936/27,028 (3.5) | 1,370/45,822 (3.0) | 8,502/27,019 (31.5) | 13,081/45,809 (28.6) | 1,646/27,039 (6.1) | 2,776/45,839 (6.1) | 2,073/27,034 (7.7) | 4,041/45,827 (8.8) |
| 30–34 | 1,957/50,915 (3.8) | 2,555/79,358 (3.2) | 18,762/50,893 (36.9) | 26,332/79,338 (33.2) | 3,360/50,937 (6.6) | 5,126/79,386 (6.5) | 4,179/50,923 (8.2) | 7,118/79,373 (9.0) |
| 35–39 | 1,126/22,983 (4.9) | 1,355/34,659 (3.9) | 10,182/22,977 (44.3) | 13,824/34,641 (39.9) | 1,933/22,994 (8.4) | 2,595/34,674 (7.5) | 2,159/22,987 (9.4) | 3,407/34,664 (9.8) |
| ≥40 | 253/3,491 (7.2) | 299/5,002 (6.0) | 1,872/3,491 (53.6) | 2,537/5,001 (50.7) | 417/3,492 (11.9) | 600/5,005 (12.0) | 430/3,492 (12.3) | 622/5,004 (12.4) |
| Total | 4,456/108,851 (4.1) | 5,882/174,067 (3.4) | 40,429/108,812 (37.2) | 57,747/174,013 (33.2) | 7,626/108,897 (7.0) | 11,755/174,136 (6.8) | 9,198/108,869 (8.4) | 16,196/174,098 (9.3) |

Hidden overweight was defined as prepregnancy BMI 23.0–24.9 kg/m$^2$, classified as normal weight under standard WHO criteria but as overweight under Asian-specific thresholds. Normal weight was defined as BMI 18.5–22.9 kg/m$^2$. Asian Indian mothers were identified using the NVSS detailed maternal race recode. Crude prevalences are unadjusted. Denominators exclude births with missing data for each outcome. The age group younger than 20 years was omitted from age-stratified display because of small cell size but is included in the total row. Abbreviations: BMI, body mass index; CD, caesarean delivery; HD, hypertensive disorders; LBW, low birthweight; PTB, preterm birth; WHO, World Health Organization..

**eTable 2.** Characteristics of birth records excluded from and included in the primary analytic sample, United States, 2016–2023.

| Characteristic | Excluded (n = 1,926,693) | Included (n = 28,024,153) | \|d\| |
|---|---|---|---|
| Maternal age, mean (SD) | 30.1 (5.9) | 29.1 (5.8) | 0.17 |
| Birthweight, g, mean (SD) | 2,806.3 (960.6) | 3,291.7 (578.9) | 0.61 |
| Caesarean delivery, % | 52.7 | 30.6 | 0.46 |
| Low birthweight, % | 33.8 | 6.6 | 0.72 |
| NH White, % | 41.5 | 51.7 | 0.20 |
| NH Asian, % | 4.9 | 6.3 | 0.06 |
| Hispanic, % | 19.8 | 24.6 | 0.12 |
| Unknown race/ethnicity, %[a] | 14.5 | 0.0 | — |

[a]Mothers with unknown race/ethnicity (coded as unknown in the NVSS file) were excluded from the analytic sample by design and therefore appear only in the excluded group.

|d|, absolute standardized difference; NH, non-Hispanic; SD, standard deviation.

**eTable 3.** Sample sizes and event counts by model, United States, 2016–2023

**Panel A.** Events and births by Asian-specific BMI category among non-Hispanic Asian mothers, used to estimate the risk ratios in Table 3 (complete-case analysis)

| BMI category (kg/m$^2$) | Hypertensive disorders, n/N (%) | Caesarean delivery, n/N (%) | Preterm birth, n/N (%) | Low birthweight, n/N (%) |
|---|---|---|---|---|
| <18.5 | 1,783/83,964 (2.12) | 19,750/83,937 (23.53) | 5,535/83,997 (6.59) | 7,702/83,980 (9.17) |
| 18.5–22.9 (reference) | 16,590/538,655 (3.08) | 151,331/538,459 (28.10) | 34,629/538,917 (6.43) | 38,098/538,822 (7.07) |
| 23.0–24.9 | 10,996/241,107 (4.56) | 79,827/241,028 (33.12) | 17,389/241,249 (7.21) | 17,423/241,184 (7.22) |
| 25.0–27.4 | 12,124/206,617 (5.87) | 74,307/206,540 (35.98) | 16,291/206,726 (7.88) | 15,695/206,661 (7.59) |
| ≥27.5 | 25,161/269,286 (9.34) | 110,066/269,189 (40.89) | 26,000/269,365 (9.65) | 22,315/269,264 (8.29) |
| Total | 66,654/1,339,629 (4.98) | 435,281/1,339,153 (32.50) | 99,844/1,340,254 (7.45) | 101,233/1,339,911 (7.56) |

**Panel B.** Births included in each model

| Analysis | Eligible births | Hypertensive disorders | Caesarean delivery | Preterm birth | Low birthweight |
|---|---|---|---|---|---|
| Asian BMI models, complete case (Table 3, Figure 2) | 1,758,116 | 1,339,629 | 1,339,153 | 1,340,254 | 1,339,911 |
| Asian BMI models, missing-indicator for marital status (eTable 6) | 1,758,116 | 1,753,843 | 1,753,367 | 1,754,491 | 1,754,133 |
| Slope-change models, BMI 18.5–27.4 (eTable 4) | 1,300,716 | 986,379 | 986,027 | 986,892 | 986,667 |

| | | | | | |
|---|---|---|---|---|---|
| Asian–White spline models (eTable 5, eFigure 2) | 16,244,194 | 14,996,621 | 14,990,774 | 15,004,335 | 15,002,321 |

N, births with complete outcome and covariate data. Most exclusions from complete-case models (418,487 of 1,758,116 Asian births, 23.8%) reflect missing marital status, which was not reported for 23.6% of Asian births. Subgroup-specific Ns are shown in Table 4.

**eTable 4.** Test for a change in slope at BMI 23.0 kg/m$^2$ among non-Hispanic Asian mothers with BMI 18.5–27.4 kg/m$^2$, United States, 2016–2023

| Outcome | N | RR per 1 kg/m$^2$, BMI <23.0 (95% CI) | RR per 1 kg/m$^2$, BMI ≥23.0 (95% CI) | Ratio of per-unit RRs (95% CI) | P |
|---|---|---|---|---|---|
| Hypertensive disorders | 986,379 | 1.13 (1.12, 1.14) | 1.12 (1.11, 1.13) | 0.991 (0.976, 1.006) | 0.24 |
| Caesarean delivery | 986,027 | 1.05 (1.04, 1.05) | 1.04 (1.03, 1.04) | 0.990 (0.985, 0.994) | <0.001 |
| Preterm birth | 986,892 | 1.01 (1.01, 1.02) | 1.04 (1.03, 1.04) | 1.025 (1.014, 1.037) | <0.001 |
| Low birthweight | 986,667 | 0.95 (0.95, 0.96) | 1.02 (1.01, 1.02) | 1.067 (1.055, 1.079) | <0.001 |

Modified Poisson regression with robust variance, with continuous BMI and a hinge term, max(BMI − 23.0, 0). Slopes were estimated on the log-risk scale. The ratio of per-unit RRs is the exponentiated hinge coefficient, equal to the RR per 1 kg/m$^2$ at or above 23.0 divided by the RR per 1 kg/m$^2$ below 23.0; a value of 1 indicates no change in slope at 23.0 kg/m$^2$. Models adjusted for maternal age, education, marital status, live birth order, prenatal care timing, payment source, smoking, nativity, birth year, infant sex, and detailed Asian subgroup. CI, confidence interval; RR, risk ratio.

**eTable 5.** Race × BMI interaction tests from restricted cubic spline models and Asian vs White risk ratios at the same BMI, United States, 2016–2023

**Panel A.** Wald chi-square tests for interaction (all P < 0.0001)

| Model | df | Hypertensive disorders | Caesarean delivery | Preterm birth | Low birthweight |
|---|---|---|---|---|---|
| Race × BMI, overall | 4 | 505.3 | 1,260.7 | 1,770.3 | 2,473.8 |
| Race × BMI, US-born mothers | 4 | 232.7 | 129.6 | 173.1 | 147.0 |
| Race × BMI, foreign-born mothers | 4 | 69.7 | 155.4 | 235.1 | 271.4 |
| Race × BMI, excluding 2020–2021 | 4 | 363.0 | 985.6 | 1,421.5 | 1,991.0 |
| Race × BMI × nativity | 4 | 124.0 | 61.5 | 25.9 | 28.6 |

**Panel B.** Adjusted RR (95% CI), non-Hispanic Asian vs non-Hispanic White mothers at the same BMI

| BMI (kg/m$^2$) | Hypertensive disorders | Caesarean delivery | Preterm birth | Low birthweight |
|---|---|---|---|---|
| 20.0 | 0.79 (0.78, 0.81) | 1.10 (1.09, 1.10) | 1.07 (1.06, 1.09) | 1.38 (1.37, 1.40) |
| 23.0 | 0.88 (0.87, 0.90) | 1.13 (1.12, 1.13) | 1.28 (1.26, 1.29) | 1.68 (1.66, 1.70) |
| 25.0 | 0.91 (0.90, 0.93) | 1.11 (1.11, 1.12) | 1.35 (1.34, 1.37) | 1.87 (1.84, 1.90) |
| 27.5 | 0.95 (0.94, 0.96) | 1.07 (1.07, 1.08) | 1.40 (1.38, 1.42) | 1.94 (1.92, 1.97) |
| 30.0 | 0.98 (0.97, 0.99) | 1.03 (1.02, 1.04) | 1.42 (1.40, 1.44) | 1.91 (1.88, 1.94) |

| | | | | |
|---|---|---|---|---|
| 35.0 | 1.02 (1.00, 1.03) | 0.98 (0.98, 0.99) | 1.43 (1.41, 1.45) | 1.86 (1.82, 1.89) |

Modified Poisson regression with robust variance. BMI was modelled with a restricted cubic spline (5 knots at the 5th, 27.5th, 50th, 72.5th, and 95th percentiles), with race × spline interaction terms. Models adjusted for maternal age, education, marital status, live birth order, prenatal care timing, payment source, smoking, nativity, birth year, and infant sex; nativity-stratified models omitted nativity. N for the overall models is given in eTable 3. CI, confidence interval; RR, risk ratio.

**eTable 6.** Sensitivity analyses and E-values for the BMI 23.0–24.9 kg/m2 risk ratio, non-Hispanic Asian mothers, United States, 2016–2023

**Panel A.** Approaches to missing marital status, adjusted RR (95% CI) for BMI 23.0–24.9 vs 18.5–22.9 kg/$m^2$

| Approach | Hypertensive disorders | Caesarean delivery | Preterm birth | Low birthweight |
|---|---|---|---|---|
| Complete case (primary) | 1.44 (1.40, 1.47) | 1.14 (1.13, 1.14) | 1.08 (1.06, 1.10) | 0.95 (0.93, 0.97) |
| Multiple imputation, 50 datasets | 1.44 (1.41, 1.47) | | | |
| Missing-indicator category | 1.44 (1.41, 1.47) | 1.13 (1.13, 1.14) | 1.08 (1.06, 1.10) | 0.95 (0.94, 0.96) |
| Marital status omitted | 1.44 (1.42, 1.47) | 1.13 (1.13, 1.14) | 1.08 (1.06, 1.10) | 0.95 (0.94, 0.97) |

**Panel B.** Hypertensive disorders, alternative definition and study period

| Analysis | N | Adjusted RR (95% CI) |
| --- | --- | --- |
| Primary definition | 1,339,629 | 1.44 (1.40, 1.47) |
| Excluding eclampsia from the outcome | 1,339,629 | 1.43 (1.40, 1.47) |
| Births in 2022–2023 only | 306,408 | 1.35 (1.29, 1.41) |

**Panel C.** E-values for hypertensive disorders

| BMI category (kg/m$^2$) | Adjusted RR (95% CI) | E-value, point estimate | E-value, lower confidence limit |
| --- | --- | --- | --- |
| 23.0–24.9 | 1.44 (1.40, 1.47) | 2.24 | 2.15 |
| ≥27.5 | 2.79 (2.73, 2.85) | 5.02 | 4.90 |

E-values were calculated on the risk ratio scale, as described in the Methods. Modified Poisson regression with robust variance, adjusted for maternal age, education, marital status (except where omitted), live birth order, prenatal care timing, payment source, smoking, nativity, birth year, infant sex, and detailed Asian subgroup. Multiple imputation used fully conditional specification (logistic regression for marital status) with 50 imputations; the fraction of missing information was <0.001. Imputation was performed for hypertensive disorders only. Marital status was missing for 23.6% of Asian births, ranging from 15.4% among Asian Indian to 33.3% among Chinese mothers. CI, confidence interval; RR, risk ratio.
